# THE EMOTIONAL IMPACT OF THE ASRM GUIDELINES ON FERTILITY PATIENTS DURING THE COVID-19 PANDEMIC

**DOI:** 10.1101/2020.03.29.20046631

**Authors:** Jenna Turocy, Alex Robles, Daniel Hercz, Mary D’Alton, Eric J. Forman, Zev Williams

## Abstract

**Objective:** To survey fertility patients’ agreement with ASRM recommendations during the COVID-19 pandemic and the emotional impact on them.

**Design:** An online survey was sent to current fertility patients

**Setting:** New York City academic fertility practice at the epicenter of the COVID-19 pandemic

**Patient(s):** Fertility patients seen within the last year

**Intervention(s):** None

**Main Outcome Measures(s):** Patient agreement with the ASRM recommendations during the COVID-19 pandemic and the emotional impact rated on a Likert scale.

**Result(s):** A total of 518 patients completed the survey for a response rate of 17%. Fifty percent of respondents had a cycle canceled due to the COVID-19 pandemic. Of those who had a cycle cancelled, 85% of respondents found it to be moderately to extremely upsetting with 22% rating it to be equivalent to the loss of a child. There was no difference on the emotional impact based on the type of cycle cancelled. Fifty-five percent of patients agreed that diagnostic procedures such as hysterosalpingograms should be cancelled while 36% of patients agreed all fertility cycles should be cancelled. Patients were slightly more likely to agree with the ASRM guidelines if they have an upcoming cycle cancelled (p = 0.041). Of all respondents 82% would have preferred to have the option to start a treatment cycle in consultation with their doctor.

**Conclusion(s):** Given the severity of the COVID-19 pandemic, the physical, financial and emotional impact of this unprecedented threat cannot be underestimated in our fertility patients.

## Introduction

The current outbreak of the novel coronavirus disease (COVID-19) has led to sweeping changes in healthcare practice and clinical recommendations (1). As the number of infected individuals requiring hospitalization increases, several medical societies have put forth guidelines to limit the strain on the healthcare system. These primarily include the suspension of non-urgent medical care and elective procedures.

On March 17^th,^ 2020, the American Society for Reproductive Medicine (ASRM) published recommendations calling for stopping the initiation of any new treatment cycles, including ovulation induction (OI), intrauterine inseminations (IUI), and in vitro fertilization (IVF) (2). In addition, ASRM recommended strong consideration should be given to suspending all embryo transfers, whether fresh or frozen. In light of these recommendations, fertility centers across the United States have had widespread cycle cancellations for patients planning or undergoing fertility treatment.

At this time, little is known on the impact of COVID-19 on pregnancy, transmission, and fetal wellbeing, but preliminary data looks promising (3-6). As more information is gathered, guidelines will likely continue to evolve. However, fertility treatment is often a time-sensitive issue, particularly for patients with advanced reproductive age or diminished ovarian reserve. According to the World Health Organization (WHO) infertility is defined as “a disease of the reproductive system defined by the failure to achieve a clinical pregnancy after 12 months or more of regular unprotected sexual intercourse (7).” The indefinite postponement of fertility treatment can lead to an irreversible ability to conceive with their own gametes in certain patient populations. According to ASRM, 74,000 babies were born from almost 280,000 ART cycles done in the U.S. in 2018 (8). Thus, the recommendations from ASRM could have a sweeping impact on a large number of patients. Furthermore, the uncertainty of this pandemic can also have a significant psychological, emotional, and financial burden on all patients, particularly as patients face the possibility of losing their insurance.

The purpose of the present study is to survey patients at a large academic fertility center in New York at the epicenter of the COVID-19 pandemic to determine the extent of the emotional and psychological impact this situation imposes.

## Materials and Methods

An 18-item survey was constructed to assess fertility patients’ reactions to the initial ASRM recommendations during the COVID-19 pandemic. The survey was sent to all patients seen at a New York academic fertility practice between December 2019 to May 2020. The survey was sent using Qualtrics platform and all respondents were anonymous with no unique identifiers collected. The survey was initially sent as a quality improvement project to guide the practice in its management of patient expectations and future treatment. The study was later approved by Columbia University Institutional Review Board for research and publication purposes.

The data were collected over a 48-hour period, including demographic characteristics of respondents such as age, sex, parity and prior fertility treatments. Patient agreement with ASRM recommendations and its emotional impact was rated on a Likert scale. Ordinal data such as responses rated on a Likert scale were analyzed using Mann-Whitney Wilcoxon testing and responses were compared using Fisher exact or chi-square test as appropriate, with significance at p<0.05.

## Results

The survey was sent to 3100 patients. A total of 518 patients, 92% female and 8% male completed the survey for a response rate of 17%. The average age was 37 (range 23-52) [Table 1]. Of the respondents, 24% had children and 66% had previously done at least a one fertility treatment previously. Of those who had completed a prior fertility treatment, 38% were intrauterine inseminations (IUI), and 39% were in vitro fertilization (IVF) and 23% were embryo transfers (ET).

**Table 1.**
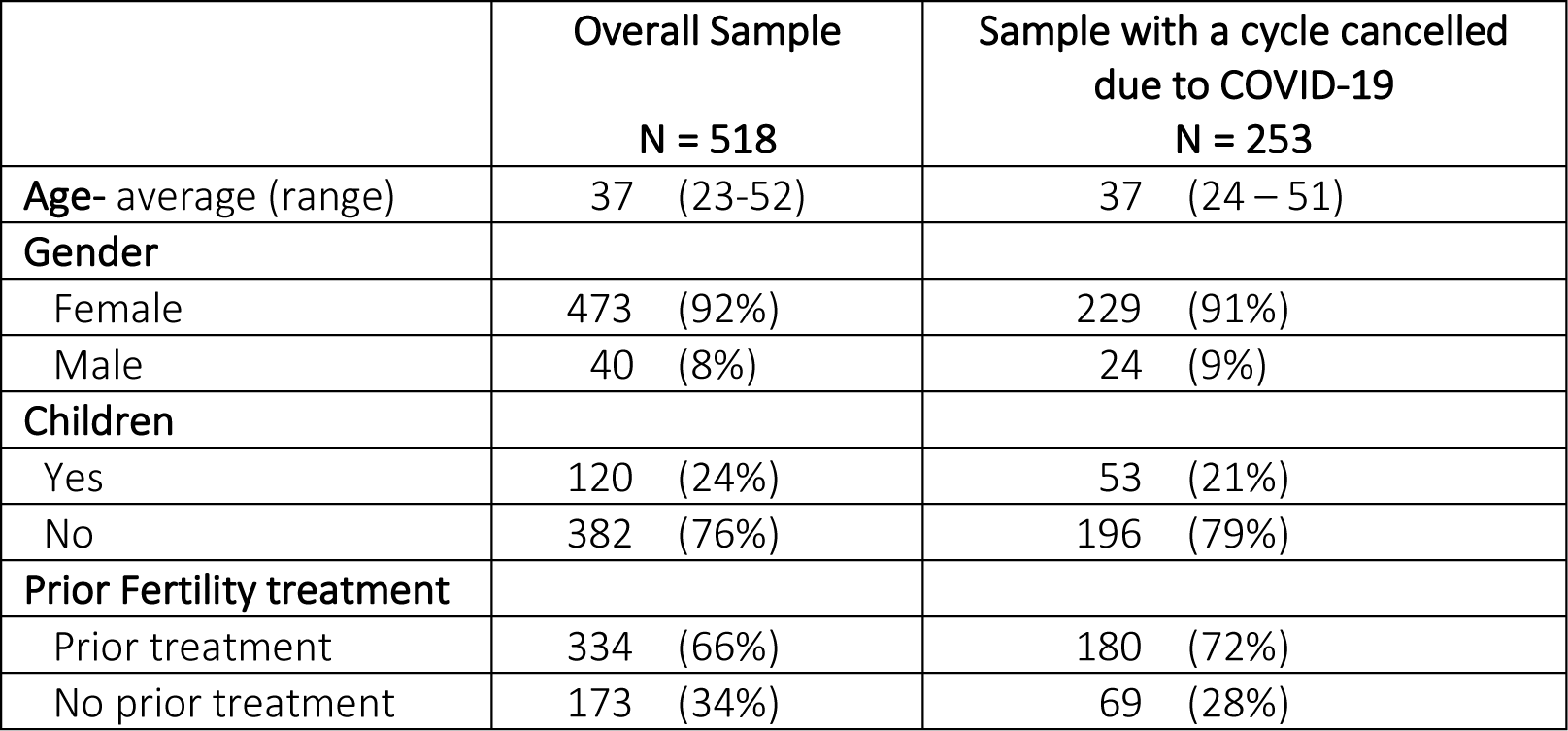
Demographic Patient Characteristics

Fifty percent of respondents had a cycle canceled due to the COVID-19 pandemic. Of those who had a cycle canceled, 5% had a timed intercourse (TIC) cycle canceled, 23% had an IUI cycle canceled, 10% had an IVF cycle with planned fresh embryo transfer cycle canceled, 27% had an IVF cycle with plan to freeze all embryos, 3% had an egg freeze cycle canceled and 30% had a frozen embryo transfer (FET) cycle canceled. Of those who had a cycle canceled, 96% found it to be upsetting and 4% found it not upsetting. 22% found it to be extremely upsetting where extremely upsetting was defined as the equivalent of the loss of a child (Figure 1). There was not a difference on the emotional impact based on the type of cycle canceled.

**Figure 1.**
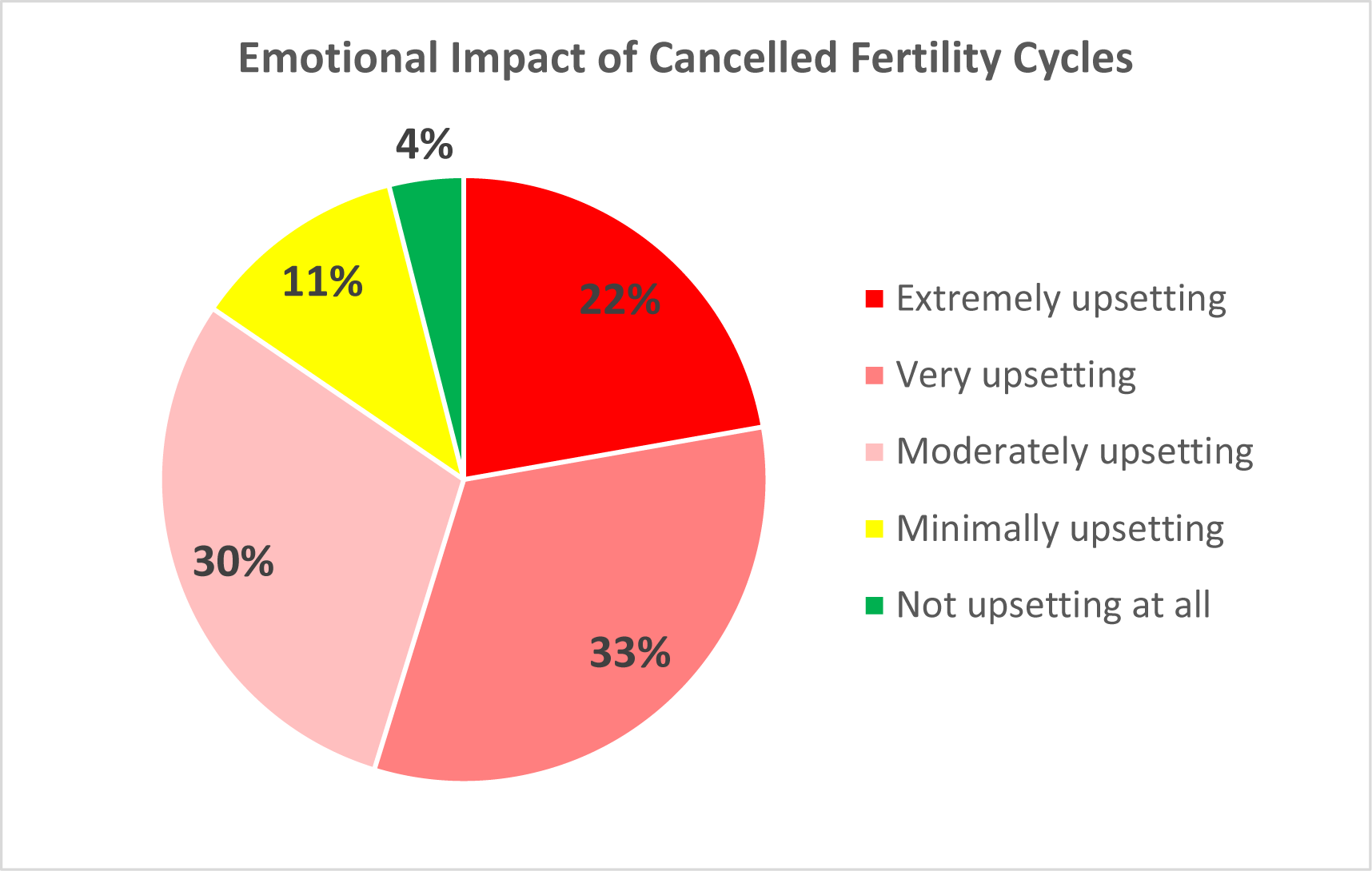
Emotional impact of cancelled fertility cycles due to COVID-19 on patients. Extremely upsetting was defined as the equivalent of the loss of a child.

The degree of agreement or disagreement of patients with the ASRM recommendations is shown in Figure 2. Fifty five percent of patients agreed that diagnostic procedures such as hysterosalpingograms should be cancelled while 36% of patients agreed all fertility cycles should be cancelled. Patients were slightly more likely to agree with ASRM guidelines if they had an upcoming cycle cancelled (P = 0.041). 40% of those patients agreed all fertility cycles should be cancelled compared to 30% without cancellations. There was not a significant difference based whether they had any children.

**Figure 2.**
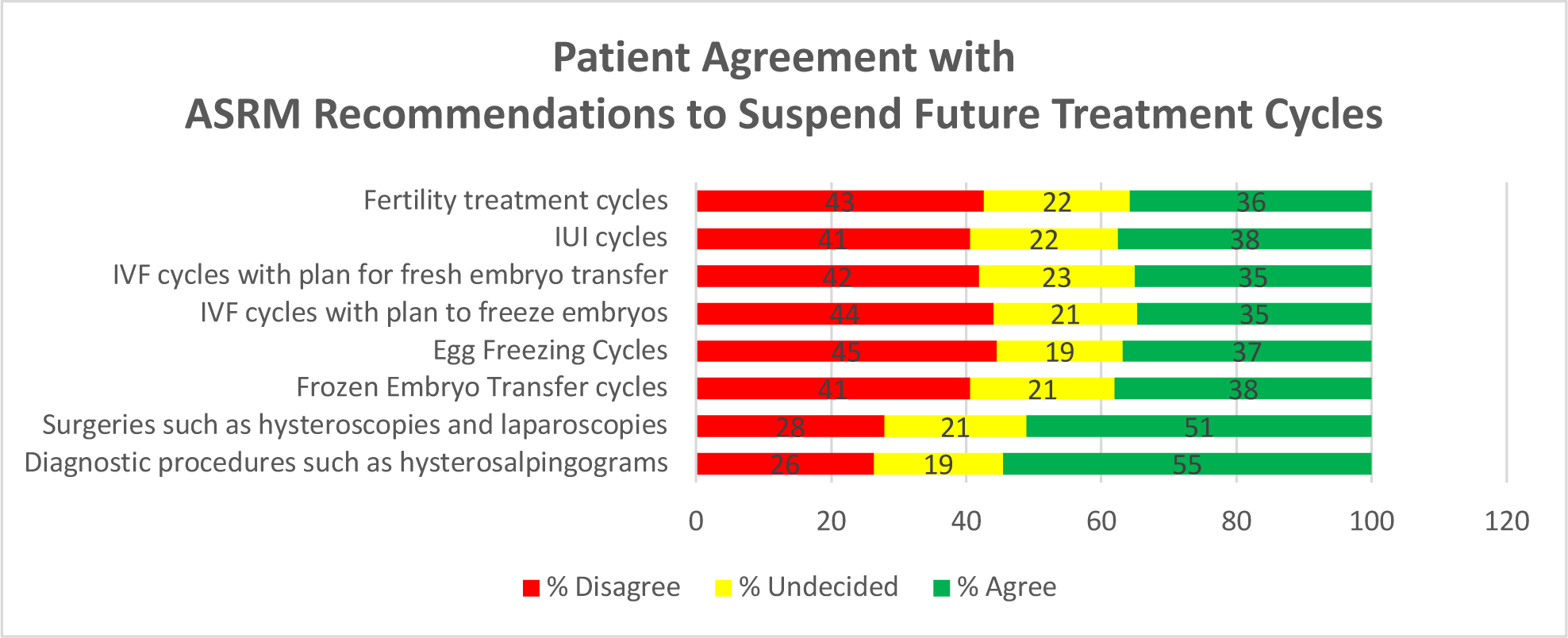
Fertility patient’s agreement or disagreement with ASRM’s recommendation to suspend the initiation of new fertility treatment cycles and procedures.

Of all respondents 82% would have preferred to have the option to start a treatment cycle in consultation with their doctor. Given an option, 52% would have chosen to start a new cycle, 24% would not have and 24% were unsure (Figure 3). Of those who had a cycle canceled 86% would have preferred to have the option to start a treatment cycle in consultation with their doctor. Given an option, 58% would have chosen to start a new cycle, 20% would not have and 21% were unsure.

**Figure 3.**
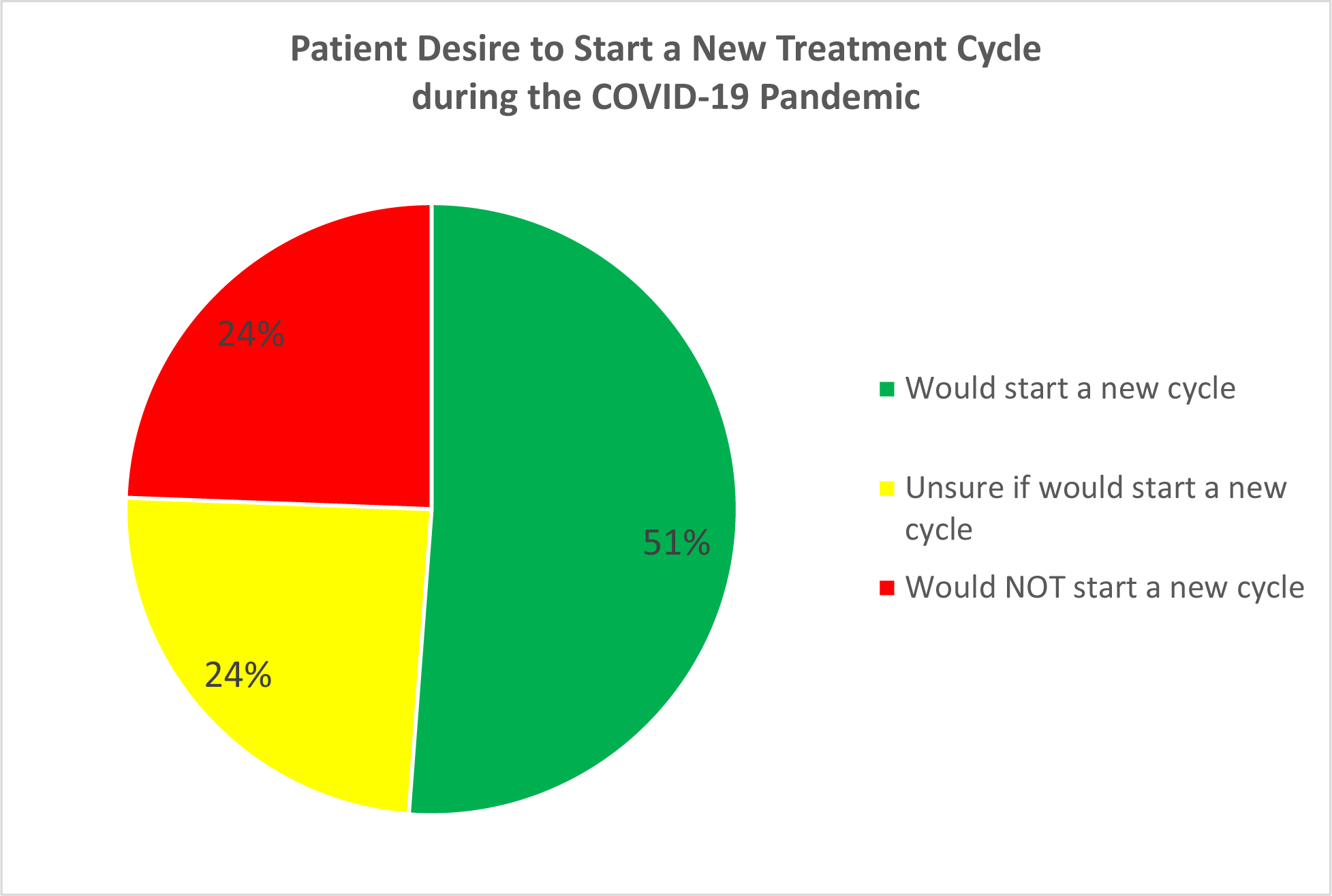
Fertility patients’ desire to start a new cycle during the COVID-19 pandemic.

## Discussion

This study provides feedback on the new ASRM guidelines from patients in what is currently the most significantly impacted area in the U.S. This information may be helpful in the context of the many considerations necessary in formulating policies to maximize our ability to care for patients, while minimizing the spread of disease and conserving healthcare resources.

Our survey was conducted in New York City, at a time when it was considered the epicenter of the pandemic and days after New York State had been placed in a “state of emergency” requiring all non-essential businesses to be closed and all elective surgical procedures to be cancelled. In addition, public health initiatives have urged all people to practice “social distancing,” limiting travel and outdoor activity in an effort to prevent, contain, and mitigate disease propagation. Despite the current situation, our survey demonstrated that 49% of respondents report that they would choose to start a new cycle under the current COVID-19 circumstances. Moreover, 85% of respondents reported having their fertility cycle cancelled was moderately to extremely upsetting, with 22% of those equating the cycle cancellation as the equivalent to losing a child. A diagnosis of infertility is associated with significant emotional and psychological consequences in both men and women (9, 10), underscoring the emotional burden that treatment cancellation can add. During this time of uncertainty and fear, the widespread cancellation of fertility treatment cycles may add to the emotional impact already inherent in the infertile population.

There are limitations to this study. First, responses from patients in other parts of the country that are not so significantly impacted from COVID-19 may differ from our data. Secondly, our response rate was 17%. Due to the anonymity of the survey we do not have information on the non-responders and do not know if they share the same opinions as our responders. It is possible that those who responded had the strongest opinions. Lastly, the situation with COVID-19 is very fluid. As we gain a deeper understanding of the long-term impact of the pandemic and see how the situation evolves on a local, national and global scale, patients’ feelings may change.

## Conclusion

Patients have a mixed opinion regarding the ASRM recommendations concerning fertility treatment at the current time of the COVID-19 epidemic and many were very upset by the cancellation of fertility treatment cycles.

## Data Availability

All data can be made available upon request

## References

1. Center for Disease Control. Interim Guidance for Healthcare Facilities: Preparing for Community Transmission of COVID-19 in the United States. Available at cdc.gov/coronavirus/2019-ncov/healthcare-facilities/guidance-hcf.html. Accessed on March 27, 2020

2. American Society for Reproductive Medicine. Patient Management and Clinical Recommendations During The Coronavirus (COVID-19) Pandemic. Available at https://www.asrm.org/news-and-publications/covid-19/statements/patient-management-and-clinical-recommendations-during-the-coronavirus-covid-19-pandemic. Accessed on March 27, 2020

3. Liu D, Li L, Wu X, Zheng D, Wang J, Yang L, et al. Pregnancy and Perinatal Outcomes of Women With Coronavirus Disease (COVID-19) Pneumonia: A Preliminary Analysis. AJR Am J Roentgenol 2020;

4. Fan C, Lei D, Fang C, Li C, Wang M, Liu Y, et al. Perinatal Transmission of COVID-19 Associated SARS-CoV-2: Should We Worry? Clin Infect Dis 2020;

5. Schwartz DA. An Analysis of 38 Pregnant Women with COVID-19, Their Newborn Infants, and Maternal-Fetal Transmission of SARS-CoV-2: Maternal Coronavirus Infections and Pregnancy Outcomes. Arch Pathol Lab Med 2020;

6. Chen H, Guo J, Wang C, Luo F, Yu X, Zhang W, et al. Clinical characteristics and intrauterine vertical transmission potential of COVID-19 infection in nine pregnant women: a retrospective review of medical records. Lancet 2020;

7. World Health Organization, “The International Committee for Monitoring Assisted Reproductive Technology (ICMART) and the World Health Organization (WHO) Revised Glossary on ART Terminology,” 2009

8. American Society for Reproductive Medicine. More than 74 Thousand Babies Born from Assisted Reproductive Technology Cycles Done in 2018. Available at https://www.asrm.org/news-and-publications/news-and-research/press-releases-and-bulletins/more-than-74-thousand-babies-born-from-assisted-reproductive-technology-cycles-done-in-2018--record-90-of-babies-born-from-art-are-singletons/. Accessed on March 27, 2020.

9. Hasanpoor-Azghdy SB, Simbar M, Vedadhir A. The emotional-psychological consequences of infertility among infertile women seeking treatment: Results of a qualitative study. Iran J Reprod Med 2014;

10. Fisher JRW, Hammarberg K. Psychological and social aspects of infertility in men: An overview of the evidence and implications for psychologically informed clinical care and future research. Asian J. Androl. 2012;

